# Clinical characteristics and factors associated with admission to intensive care units in hospitalized COVID-19 patients in Lyon University Hospitals, France

**DOI:** 10.1101/2020.06.09.20125286

**Authors:** Philippe Vanhems, Marie-Paule Gustin, Christelle Elias, Laetitia Henaff, Cédric Dananché, Beatrice Grisi, Elodie Marion, Nagham Khanafer, Delphine Hilliquin, Sophie Gardes, Solweig Gerbier-Colomban, Selilah Amour, Elisabetta Kuczewski, Vanessa Escuret, Bruno Lina, Mitra Saadatian-Elahi

**Author notes:** On behalf of COVID-Outcomes-HCL Consortium Laurent Argaud, Frédéric Aubrun, Marc Bonnefoy, Maude Bouscambert-Duchamp, Roland Chapurlat, Dominique Chassard, Christian Chidiac, Michel Chuzeville, Cyrille Confavreux, Sébastien Couraud, Gilles Devouassoux, Isabelle Durieu, Michel Fessy, Sylvain Gaujard, Alexandre Gaymard, Arnaud Hot, Géraldine Martin Gaujard, Emmanuel Morelon, Vincent Piriou, Véronique Potinet, Jean-Christophe Richard, Thomas Rimmele, Pascal Sève, Alain Sigal, Karim Tazarourte. **Correspondence to** Professor Philippe Vanhems, Groupement Hospitalier Centre, Unité d’Hygiène, Epidémiologie, Prévention - Bâtiment 1, 5, place d’Arsonval-69437 Lyon cedex 3. **Author contribution** PV designed the study. MPG, CE, LH, CD and MSE participated in the design of the CRF. LH, BG, SG, SGC, SA and EK completed the data entry. VE and BL participated in the laboratory analysis. PV, MPG and CE performed the statistical analyses. PV, MPG, CE, CD, EM, NK, DH and MSE reviewed and interpreted the statistical analyses. PV, MPG, CE and MSE drafted the manuscript. All co-authors reviewed and approved the final version of the manuscript.

## Abstract

**Introduction:** A new respiratory virus, SARS-CoV-2, has emerged and spread worldwide since late 2019. This study aims at analyzing clinical presentation on admission and the determinants associated with direct admission or transfer to intensive care units (ICUs) in hospitalized COVID-19 patients.

**Patients and Methods:** In this prospective hospital-based study, socio-demographic, clinical and biological characteristics, on admission, of adult COVID-19 hospitalized patients were prospectively collected and analyzed. The outcome was admission/transfer to intensive care units compared with total hospital stay in medical wards according to patient characteristics.

**Results:** Of the 412 patients included, 325 were discharged and 87 died in hospital. Multivariable regression showed increasing odds of admission/transfer to ICUs with male gender (OR, 1.99 [95%CI, 1.07-3.73]), temperature (OR, 1.37 [95% CI, 1.01-1.88] per degree Celsius increase), abnormal lung auscultation on admission (OR, 2.62 [95% CI, 1.40-4.90]), elevated level of CRP (OR, 6.96 [95% CI, 1.45-33.35 for CRP>100mg/L vs CRP<10mg/L). Increased time was observed between symptom onset and hospital admission (OR, 4.82 [95% CI, 1.61-14.43] for time >10 days vs time <3 days) and monocytopenia (OR, 2.49 [95% CI, 1.29-4.82]). Monocytosis was associated with lower risk of admission/transfer to ICUs (OR, 0.25 [95% CI, 0.05-1.13]).

**Conclusions:** Clinical and biological features on admission and time until admission were associated with admission to ICUs. Signs to predict worsening on admission could be partially associated with the time until admission. This finding reinforces the need for appropriate guidelines to manage COVID-19 patients in this time window.

## Introduction

Severe acute respiratory syndrome coronavirus 2 (SARS-CoV-2), first detected in December 2019 in the Hubei province of China [1–3], was declared as a pandemic by the World Health Organization on March 11, 2020. Coronavirus disease 2019 (COVID-19) is the emerging infectious disease due to SARS-CoV-2, mainly associated with lower respiratory infection even though less typical clinical features or asymptomatic cases have also been reported [4, 5]. The crude case fatality rate ranges from 2% to 4% but can reach 12% to 15% in the elderly [6].

The first cases of SARS-CoV2 infection in Europe were travellers from Wuhan who were tested positive in France (two in Paris and one in Bordeaux) on January 24, 2020 [7]. As of May 3, 2020, 131,287 confirmed cases have been reported in France, including 3,918 patients still hospitalized in intensive care units (ICUs) and 15,583 deaths in hospitals [8]. The Auvergne-Rhône-Alpes region located in the southeast of France has a population of more than 6 million inhabitants. The first confirmed case of SARS-CoV-2 in this region was identified on February 8^th^ and had reached 8,431 cases and 1,192 deaths by April 24^th^.

COVID-19 related complications, patient outcomes and mortality rates reported so far have varied considerably between countries most probably owing to differences in healthcare systems and the availability of ICU beds. Moreover, the prevalence of underlying chronic diseases such as obesity and diabetes, known to be important determinants in the clinical course and outcome of COVID-19 [9] are also different throughout the world. In addition, a large number of published reports so far have described hospitalized COVID-19 patients with incomplete data vis-à-vis hospital follow-up because a substantial proportion of patients remained hospitalized at the time of manuscript submission or publication.

Knowledge of the baseline characteristics and outcomes of hospitalized COVID-19 patients from different parts of the world is crucial for the decision-making process at national and international levels in order to properly respond to the pandemic.

The aim of this study was to report the clinical features and outcomes of confirmed COVID-19 patients admitted to Lyon university-affiliated hospitals with complete documentation of the hospital stay from February 8 to April 24, 2020 and to identify the demographic, clinical and biological characteristics on admission associated with the risk of direct admission or subsequent transfer to ICUs.

## Methods

### Study design and participants

This prospective, observational, hospital-based study (NOSO-COR, ClinicalTrials: NCT04290780) is an ongoing international multicenter study carried out in France and hospitals affiliated with the GABRIEL network [10]. However, the present paper was limited to community-acquired COVID-19 confirmed patients admitted to the four university-affiliated hospitals in Lyon (Hospices Civils de Lyon, 5,300 beds).

Any adult patient who presented an infectious syndrome based on the WHO definition of COVID-19 [11], admitted to one of the four participating university-affiliated hospitals in Lyon between February 8 and April 24, 2020, and hospitalized for a period of at least 24 hours, was included.

The study was approved by the clinical research and ethics committee of Ile de France V on March 8, 2020 (No. 20.02.27.69817 Cat 3).

### Data collection

Identification of community-acquired confirmed SARS-CoV-2 patients was based on a daily extraction of real-time reverse transcriptase-polymerase chain reaction (RT-PCR) positive patients from the virology laboratory. Electronic medical records were the main source of data collection. Demographic characteristics, underlying comorbidities, clinical, and biological parameters and patient outcome data were collected prospectively on an electronic case-report form designed especially for the purpose of the project. Clinical outcomes were monitored up to hospital discharge or death. All data were double-checked after computerization.

Nasopharyngeal swab samples were collected as part of the standard care in patients presenting signs and symptoms of SARS-CoV-2 infection. Samples were transferred to the French national reference center of respiratory viruses for the detection of SARS-CoV2 by RT-PCR [12]. Patients with positive RT-PCR results were defined as laboratory-confirmed SARS-CoV-2.

### Statistical analysis

Given the descriptive nature of this observational study and the emergency context, no statistical sample size calculation was performed. Sample size was equal to the number of patients included during the study period.

Continuous variables were reported as median and interquartile range (IQR). Categorical variables were described as frequencies (%). Comparisons between groups were performed using Mann-Whitney or Kruskall-Wallis test and Chi-square or Fisher exact test when appropriate.

Univariate logistic regression was first used to identify patient determinants known on admission that were associated with the outcome i.e. admission or transfer to an ICU versus total hospital stay in a medical ward. The determinants that were significant at 0.15 levels in univariate analysis where retained for multivariate analysis. Gender, temperature and the time between the onset of symptoms and hospital admission were first introduced in the multivariable model and then added sequentially to the model. The logistic regression model was applied to 374 patients for whom complete biological data (white blood cells, neutrophil, lymphocyte, monocyte, creatinine, red blood cells, hemoglobin and C-reactive protein) were available. Statistical tests were 2-tailed with a level of statistical significance of <.05. Statistical analysis was performed using R language version 3.5.2 (https://cran.r-project.org/).

## Results

### Patient characteristics on admission

From February 8 to April 24, 2020, a total of 412 SARS-CoV-2 confirmed patients with known date of hospital discharge or death were included. Overall, 66 patients (16.0%) were admitted directly to ICUs and 320 (77.7%) were hospitalized in medical wards, of whom 26/320 (8.1%) required subsequent transfer to ICUs. Demographic data, clinical signs and symptoms on admission for the overall study population and according to hospitalization ward are summarized in Table 1. Median age was 72.0 years [IQR, 57-83] and 56.3% were men. A total of 188 (45.6%) patients were younger than 70 years, 139 (33.7%) were aged between 70 and 84 years old and 82 (20.6%) were older than 85 years. One or more pre-existing comorbidities were present in 286 patients (69.4%): cardiovascular diseases (47.6%), diabetes (19.9%) and chronic lung diseases (15.0%) being the most common. The most frequently reported signs and symptoms on admission were cough (73.5%), dyspnea/tachypnea (64.3%), general weakness (61.4%) and fever (>37.8°C, 57.0%). Abnormal lung auscultation was observed in 229 patients (55.6%).

**Table 1.**
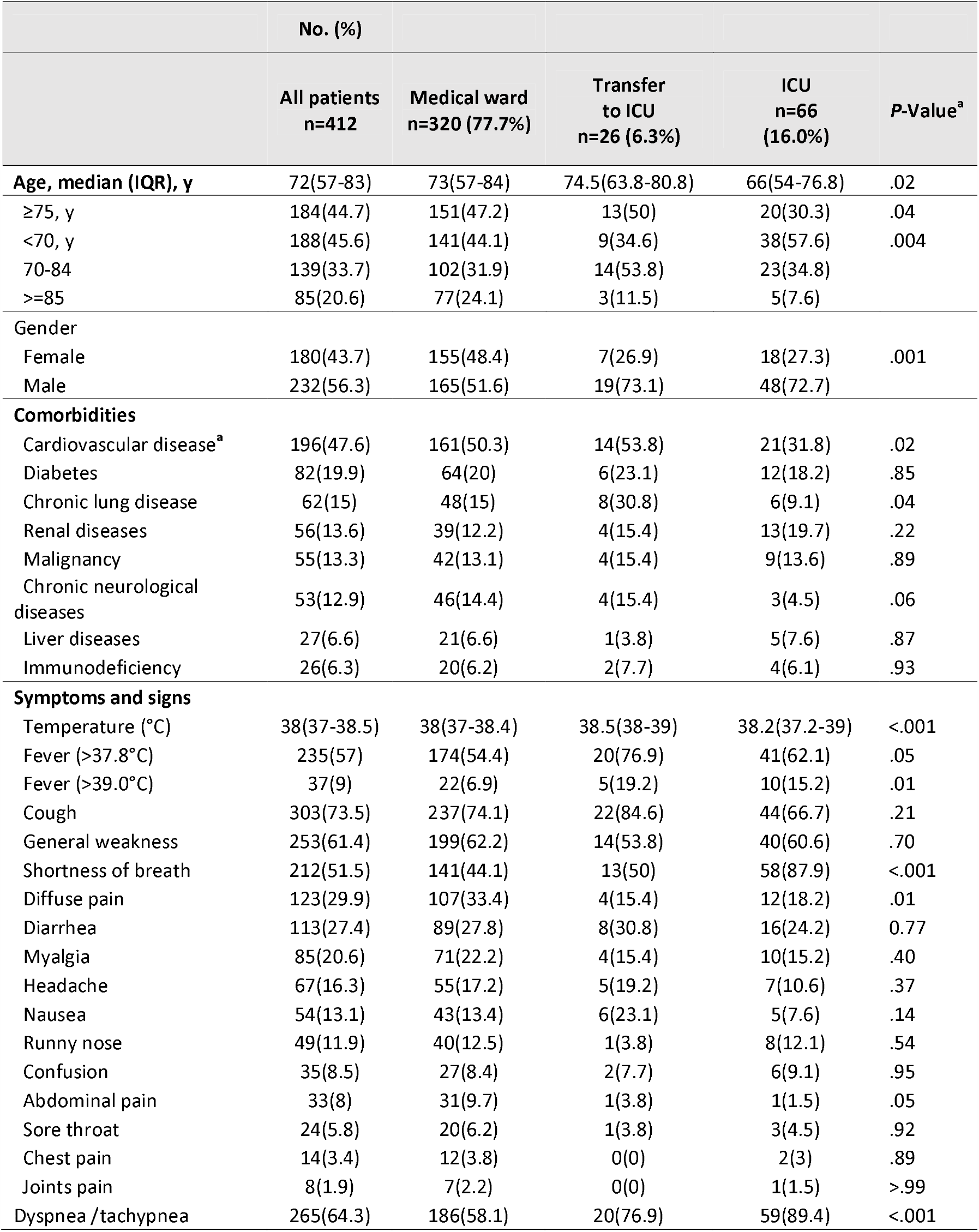
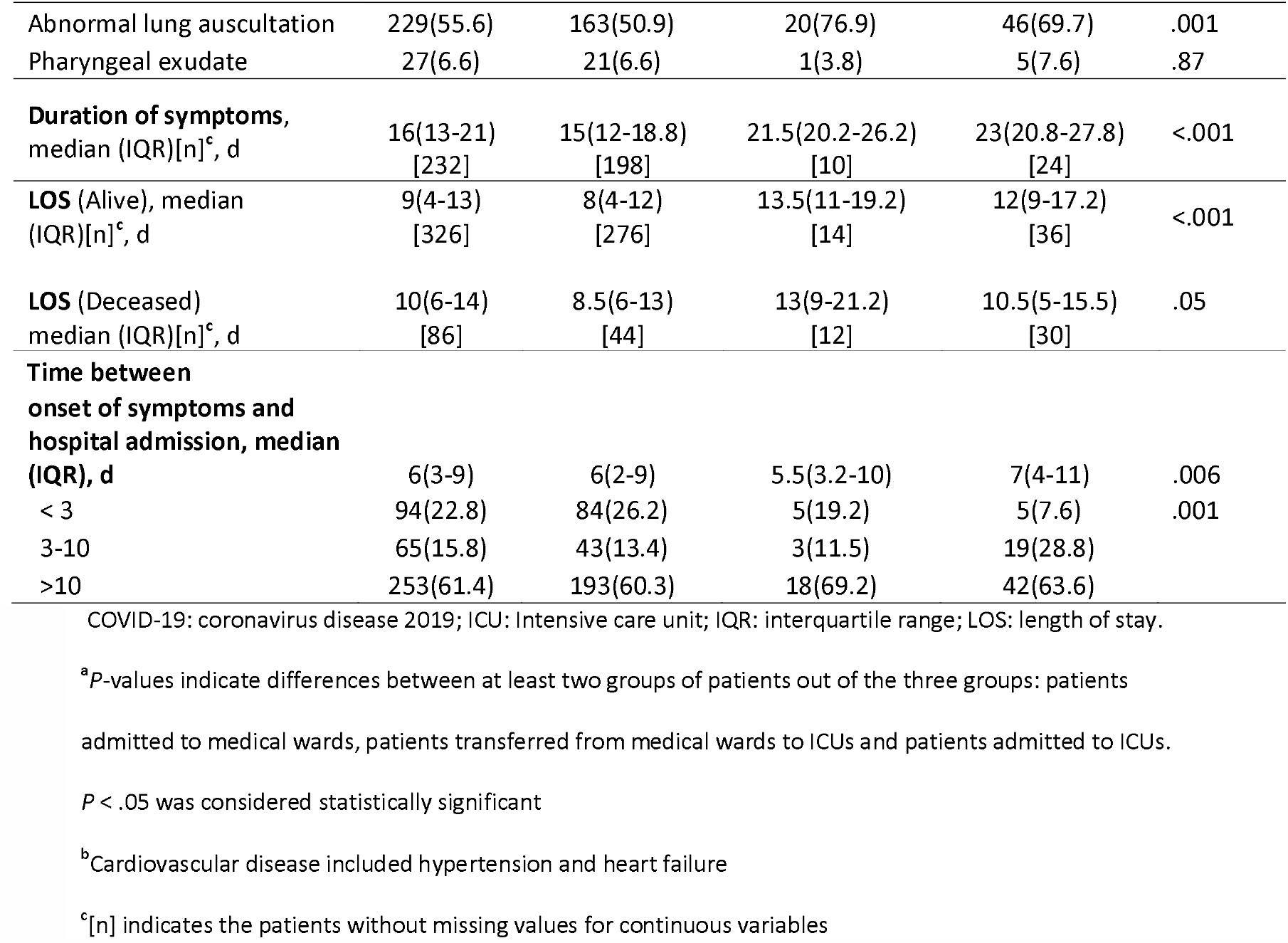
Demographic and clinical characteristics on admission of 412 confirmed COVID-19 hospitalized patients at Lyon University Hospitals, France

Males were significantly more prone to hospitalization in ICUs (*P* = .001). The proportion of patients with comorbidities was similarly distributed among medical wards and ICUs except for cardiovascular diseases (*P* = .02) and chronic lung diseases (*P* = .04). Diffuse or abdominal pain were reported significantly more often in patients hospitalized in medical wards (*P* = .01 and *P* = .05 respectively). Patients admitted or transferred to ICUs presented more often with fever (>37.8°C, 62.1% and 76.1% vs 54.4%; *P* = .05), shortness of breath (87.8% vs 44.1% and 50%; *P* < .001), showed more frequently abnormal lung auscultation (69.7% and 76.9% vs 50.9%; *P* = .002) and suffered from dyspnea/tachypnea (89.4% and 76.9% vs 58.1%; *P* < .001). In 232 patients, the duration of symptoms ((median, 23 [IQR, 20.8-27.8]; *P*<.001) was significantly higher in those admitted to ICUs while the time between symptom onset and hospital admission was significantly lower in patients hospitalized in medical wards (median, 6 [IQR, 2-9] and 5.5[IQR,3.2-10] vs 7[IQR,4-11]; *P* = .006).

As of April 24, 2020, 87 (21.1%) patients had died. The crude case fatality rate differed between patients hospitalized in medical wards (14.1%), and those directly admitted to ICUs (45.5%) or transferred to ICUs (46.2%) (*P* < .0001).

Initial biological data on admission are represented in Table 2. The majority of biological parameters were in the normal range although their values differed between patients admitted to ICUs and those hospitalized in medical wards. We noted that half of the population had marked lymphopenia, in particular, ICU patients (Median 0.7, [IQR, 0.6-1.1]; *P* = .0004). Elevated levels of lactate dehydrogenase (LDH) (N=134, Median 330.5, [IQR, 236.5-419.6]) and C-reactive protein (CRP) (N=378, median 60.7, [IQR, 23.4-122.3]) in half of the study population on admission were also noted. Analysis by hospital ward showed that the highest values of these parameters were found in patients directly admitted to ICUs.

**Table 2.**
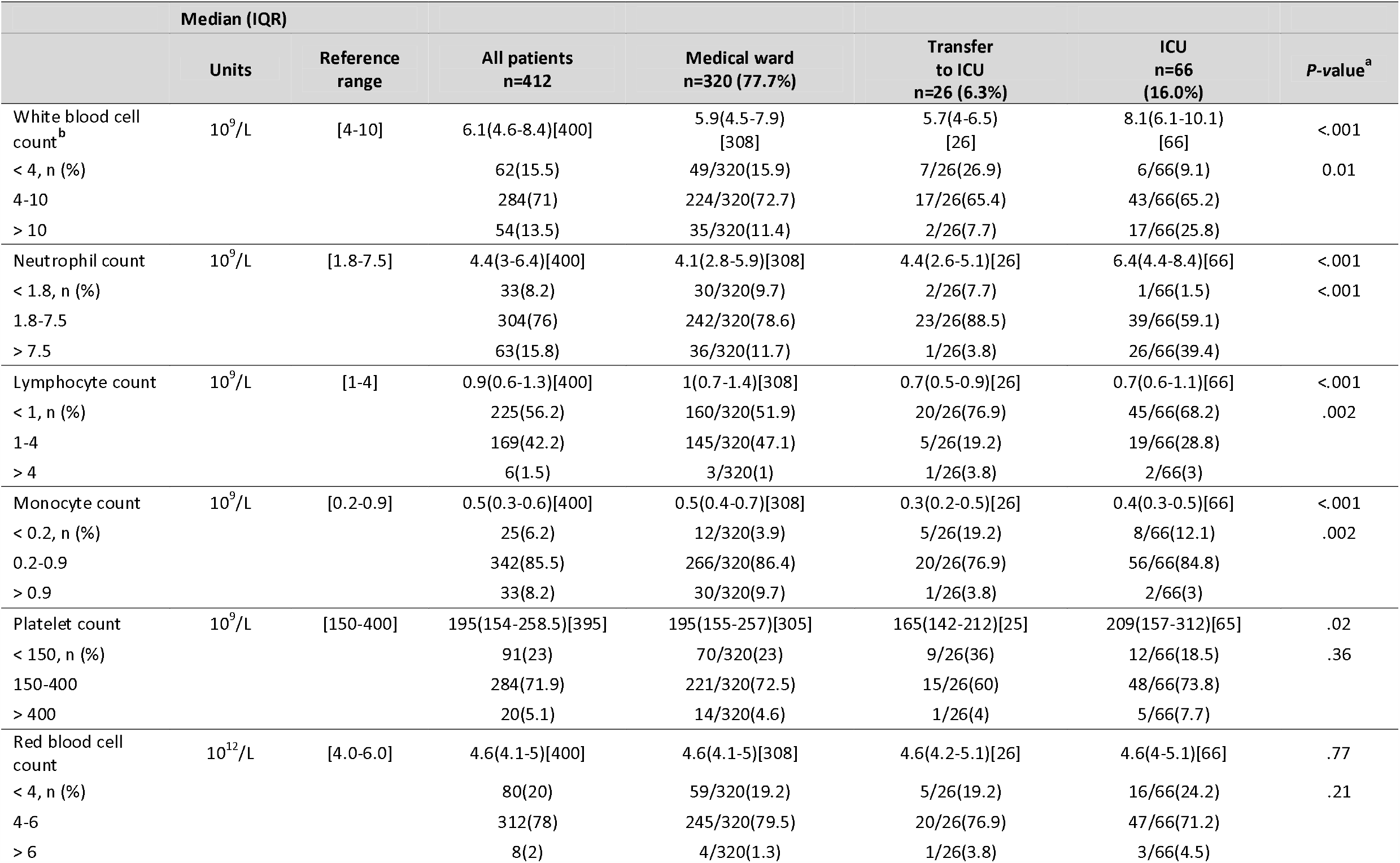
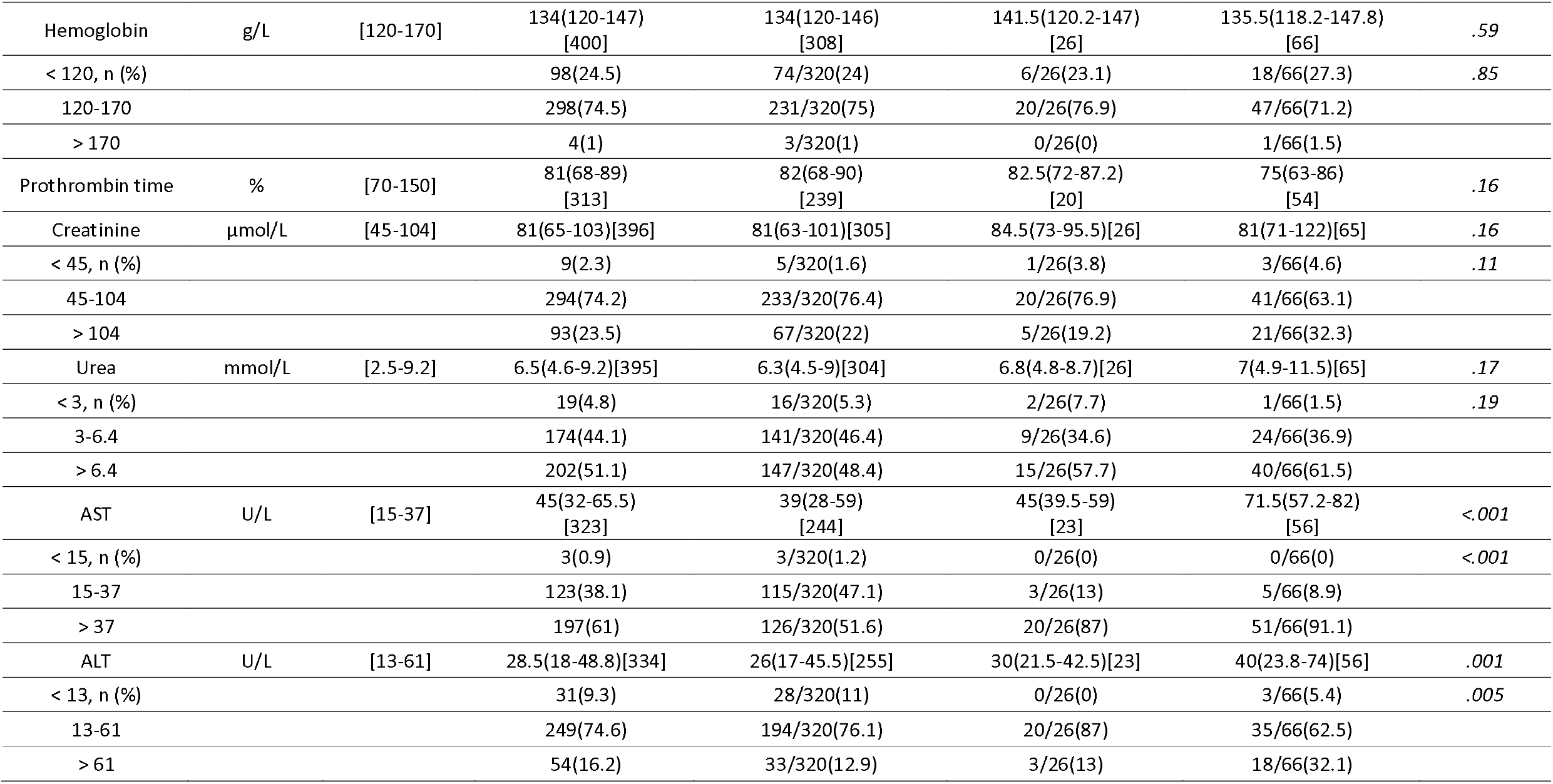
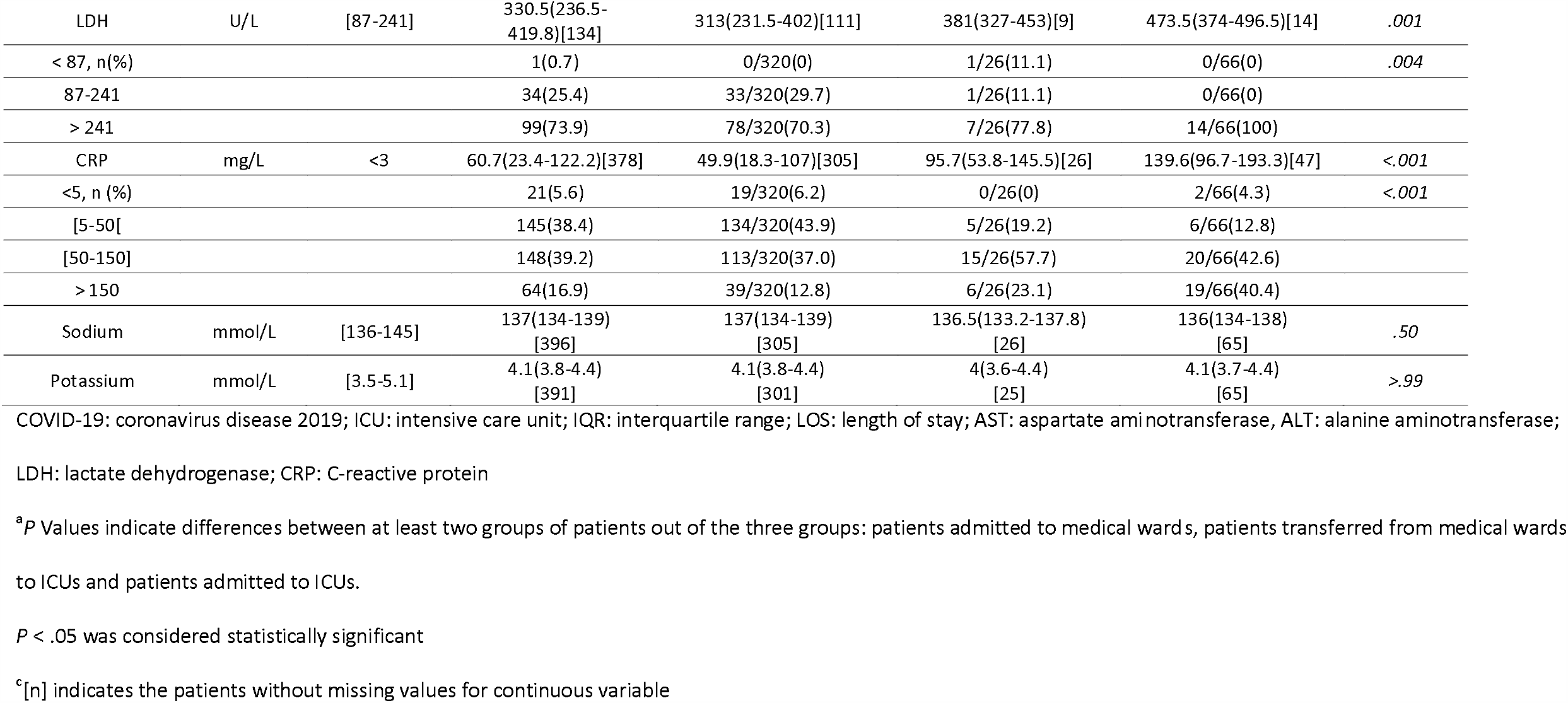
Laboratory measures on admission of confirmed Covid-19 hospitalized patients at Lyon University Hospitals, France

### Patient determinants associated with direct admission/transfer to ICUs

The analysis was based on 374 patients with complete biological data. The crude case fatality rate was not statistically different between included patients and excluded patients (20.0% (75/374) vs 31.6%, (12/38); *P* = .15).

Demographic, clinical and biological characteristics on admission associated with direct admission or subsequent transfer to ICUs are summarized in Table 3. Men had a twice higher risk of being hospitalized/transferred to ICUs (OR, 1.99 [95%CI, 1.07-3.73]; *P* = .03). Temperature (OR, 1.37 [95%CI, 1.01-1.88] per degree Celsius increase; *P* = .05) and abnormal lung auscultation on admission (OR, 2.62 [95%CI, 1.40-4.90]; *P* = .003) were associated with a higher risk of admission/transfer to ICUs. The odds of admission/transfer to ICUs revealed a statistically significant increasing trend with an elevated level of CRP (OR, 6.96 [95%CI, 1.45-33.35 for CRP> 100mg/L vs CRP < 10mg/L; *P* = .02). Increased time from symptom onset to hospital admission was also significantly associated with ICU hospitalization/transfer to ICUs (OR, 4.82 [95%CI, 1.61-14.43] for a time > 10 days vs a time < 3 days; *P* = .0053). Monocytopenia was associated with increased risk of ICU hospitalization (OR, 2.49 [95%CI, 1.29-4.82] *P* = .007) while monocytosis was associated with a lower risk of admission/transfer to ICUs (OR, 0.25 [95%CI, 0.05-1.13]; *P* = .005).

**Table 3:** Determinants associated with admission to intensive care units in 374 Covid-19 patients at Lyon University hospitals, France

|  | Adjusted OR | 95%CI | P-value |
| --- | --- | --- | --- |
| **Sex** |  |  |  |
| Female | 1(ref) |  |  |
| Male | 1.99 | 1.07-3.72 | .03 |
| **Temperature, C<sup>a</sup>** | 1.37 | 1.01-1.88 | .05 |
| **Abnormal lung auscultation** |  |  |  |
| No | 1(ref) |  |  |
| Yes | 2.62 | 1.40-4.90 | .003 |
| **C Reactive Protein, mg/L<sup>b</sup>**<br>(Normal range <5 mg/L) |  |  |  |
| < 10 | 1(ref) |  |  |
| 10-100 | 2.23 | 0.46-10.85 | .32 |
| > 100 | 6.96 | 1.45-33.35 | .02 |
| **Monocyte count, x10<sup>9</sup>/L<sup>b</sup>**<br>(Normal range 0.2-0.9 x10<sup>9</sup>/L) |  |  |  |
| 0.3-0.8 | 1(ref) |  |  |
| < 0.3 | 2.49 | 1.29-4.82 | .007 |
| > 0.8 | 0.25 | 0.05-1.13 | .07 |
| **Onset of symptoms and hospital admission, d** |  |  |  |
| < 3 | 1(ref) |  |  |
| 3 - 10 | 2.67 | 1.02-6.97 | .05 |
| > 10 | 4.82 | 1.61-14.43 | .005 |

OR: odds ratio, CI: confident interval

In multivariable logistic regression: i) ICU admission was the dependant variable and ii) sex, continuous temperature, abnormal lung auscultation, C-reactive protein, monocyte count, time between the onset of symptoùs and hospital admission discretized into 3 categories were additive independent variables, Akaike information criterion, (AIC) = 307.7, test of Hosmer and Lemeshow goodness of fit with 20 bins: *P* = 0.26

^a^odds ratio of ICU admission was multiplied by 1.37 per degree celsius increase

^b^For laboratory variables, limits of the normal range could have been changed to have at least 10% (/374) of observations in each category after variable discretization.

## Discussion and conclusions

To the best of our knowledge, this report is the largest series of hospitalized COVID-19 patients with complete follow-up data in France and complete epidemiological information already available from other European countries [13–15]. Overall, 16.6% of the patients were directly admitted to ICUs and 6.3% were transferred to ICUs from medical wards. The study comprised 412 patients with 87 deaths and 325 patients discharged alive.

The observed case fatality rate of 21.1% in this series is higher than those reported in China [16, 17], similar to what has been observed in New York City [18] and lower than the mortality rates of 26% reported in ICU-hospitalized patients in Lombardy [13]. The relatively younger age of patients in the Chinese studies (median ages: 56 and 49 respectively) could explain the lower mortality rates reported in these studies.

The most commonly known manifestations of the disease i.e. cough, weakness and fever on admission in our patients were in general similar to those reported in other studies [16, 19, 20]. As reported earlier, cardiovascular diseases and diabetes were the most common comorbidities [17, 21].

In agreement with the results of a recent single-arm meta-analysis [22], men accounted for a higher proportion of COVID-19 patients than women in the present study. Similar findings have been reported for MERS-CoV [23]. Women and men traditionally differ in their perceptions of risk [24]. In women, better adoption of protective behavior such as hand-washing [25], in particular in the context of a pandemic [26], could at least in part explain the observed results.

Consistent with respiratory viral infections, our hospitalized patients had lymphopenia and elevated levels of LDH and CRP. Lymphopenia and increased levels of LDH and CRP were also reported in the meta-analysis of 1994 COVID-19 patients [20].

Few studies have evaluated clinical characteristics on admission that could predict direct hospitalization or subsequent transfer to ICUs. Our results suggest that compared with non-ICU patients, male gender, temperature, abnormal lung auscultation on admission, high levels of CRP, monocytopenia and increased time between onset of symptoms and hospital admission could increase the risk of admission/transfer to ICUs. A high level of CRP has been reported to be an independent risk factor to assess the severity of COVID-19 [27].

The time between the onset of symptoms and hospital admission was strongly associated with ICU admission and could be influenced by multiple determinants such as age, socio-economic status, personal risk perception, access to care, geographic factors, and tools used by physicians for remote management of patients. The time between the onset of symptoms and hospital admission was inversely associated with age in our study with a shorter time for older patients (*p* < 0.05) (Supplementary material). This correlation could explain the lack of independent association between age and outcome when age was introduced after the time in the multivariable model. The time to admission could be reduced through public health messages and appropriate guidelines. In addition, we can speculate that this time also provides opportunities for inter-human transmission at the early stages of the disease before hospitalization [19].

The prospective design, inclusion of both severe and non-severe cases and complete follow-up of the study population are the main strengths of the present study. Multivariable analysis was based on 374 patients with complete biological data. However, selection bias remains low since the case fatality rate did not differ between patients who were included and not included in the model. Only biological measures on admission were analyzed because repeated measurements were most likely only performed in more severe cases.

In conclusion, even if early clinical and pathogenic events of the SARS-CoV-2 infection contribute to the prognosis, the time between the onset of symptoms and hospital admission should be considered as warning markers for more severe pathogenic processes associated with the spectrum of outcomes. This time could be reduced through appropriate preventive measures.

## Data Availability

All relevant data are within the manuscript and its Supporting Information files.

## Acknowledgements

The authors express their gratitude to 1) the Department of Health Data of Lyon Hospital: Pr. A. Duclos, Pr. F. Gueyffier, S. Vautier and M. Hervé for the creation and management of e-CRF, 2) Clinical research associates for data collection and data entry: V. Artizzu, L. Bissuel, S. Bennina, L. Dehina-Khenniche, A. Darrin, M. Grange, B. Robin, 3) staff of the virology laboratory of the Lyon hospital: Claire Bandolo, Genevieve Billaud, Maude Bouscambert-Duchamp, Emilie Frobert, Alexandre Gaymard, Laurence Josset, Christophe Ramiere, Isabelle Schuffenecker, Solange Telusson, Martine Valette, Florence Morfin for providing the results of RT-PCR tests.

## Supporting information

Distribution of the time between the onset of symptoms and hospital admission by age category

## References

[1] Hui DS, I Azhar E, Madani TA, Ntoumi F, Kock R, Dar O, et al. The continuing 2019-nCoV epidemic threat of novel coronaviruses to global health - The latest 2019 novel coronavirus outbreak in Wuhan, China. Int J Infect Dis. 2020; 91:264–266. doi: 10.1016/j.ijid.2020.01.009.

[2] Zhu N, Zhang D, Wang W, Li X, Yang B, Song J, et al. A Novel Coronavirus from Patients with Pneumonia in China, 2019. N Engl J Med. 2020;382(8):727–733. doi:10.1056/NEJMoa2001017.

[3] Lu H, Stratton CW, Tang YW. Outbreak of pneumonia of unknown etiology in Wuhan, China: The mystery and the miracle. J Med Virol. 2020;92(4):401–402. doi:10.1002/jmv.25678.

[4] Lupia T, Scabini S, Mornese Pinna S, Di Perri G, De Rosa FG, Corcione S. 2019 novel coronavirus (2019-nCoV) outbreak: A new challenge [published online ahead of print, 2020 Mar 7]. J Glob Antimicrob Resist. 2020; 21:22–27. doi: 10.1016/j.jgar.2020.02.021.

[5] Hu Z, Song C, Xu C, Jin G, Chen Y, Xu X, et al. Clinical characteristics of 24 asymptomatic infections with COVID-19 screened among close contacts in Nanjing, China. Sci China Life Sci. 2020;63(5):706–711. doi:10.1007/s11427-020-1661-4.

[6] Perlman S. Another Decade, Another Coronavirus. N Engl J Med. 2020;382(8):760–762. doi:10.1056/NEJMe2001126.

[7] Bernard Stoecklin S, Rolland P, Silue Y, Mailles A, Campese C, Simondon A, et al. First cases of coronavirus disease 2019 (COVID-19) in France: surveillance, investigations and control measures, January 2020. Euro Surveill. 2020;25(6):2000094. doi:10.2807/1560-7917.ES.2020.25.6.2000094.

[8] Santé Publique France. http://www.santepubliquefrance.fr/maladies-et- www.santepubliquefrance.fr/maladies-et-traumatismes/maladies-et-infections-respiratoires/infection-a-coronavirus/articles/infection-au-nouveau-coronavirus-sars-cov-2-covid-19-france-et-monde#block-242818. (Accessed on May 4 2020)

[9] Simonnet A, Chetboun M, Poissy J, Raverdy V, Noulette J, Duhamel A, et al. High prevalence of obesity in severe acute respiratory syndrome coronavirus-2 (SARS-CoV-2) requiring invasive mechanical ventilation [published online ahead of print, 2020 Apr 9]. Obesity (Silver Spring). 2020;10.1002/oby.22831. doi:10.1002/oby.22831.

[10] Saadatian-Elahi M, Picot-Sanchez V, Hénaff L, Pradel F, Escuret V, Dananché C, et al. MedRxiv 2020.04.08.20057471; doi: https://doi.org/10.1101/2020.04.08.20057471.

[11] World Health Organisation. Coronavirus disease 2019 (COVID-19) Situation Report – 70. Page 9. https://www.who.int/docs/default-source/coronaviruse/situation-reports/20200330-sitrep-70-covid-19.pdf?sfvrsn=7e0fe3f8_2.

[12] Corman VM, Landt O, Kaiser M, Molenkamp R, Meijer A, Chu DK, et al. Detection of 2019 novel coronavirus (2019-nCoV) by real-time RT-PCR. Euro Surveill. 2020;25(3):2000045. doi:10.2807/1560-7917.ES.2020.25.3.2000045.

[13] Grasselli G, Zangrillo A, Zanella A, Antonelli M, Cabrini L, Castelli A, et al. Baseline Characteristics and Outcomes of 1591 Patients Infected With SARS-CoV-2 Admitted to ICUs of the Lombardy Region, Italy [published online ahead of print, 2020 Apr 6]. JAMA. 2020;323(16):1574–1581. doi:10.1001/jama.2020.5394.

[14] Spiteri G, Fielding J, Diercke M, Campese C, Enouf V, Gaymard A, et al. First cases of coronavirus disease 2019 (COVID-19) in the WHO European Region, 24 January to 21 February 2020. Euro Surveill. 2020;25(9):2000178. doi:10.2807/1560-7917.ES.2020.25.9.2000178.

[15] Hodcroft EB. Preliminary case report on the SARS-CoV-2 cluster in the UK, France, and Spain. Swiss Med Wkly. 2020;150(9-10):10.4414/smw.2020.20212. Published 2020 Feb 27. doi:10.4414/smw.2020.20212

[16] Wang D, Hu B, Hu C, Zhu F, Liu X, Zhang J, et al. Clinical Characteristics of 138 Hospitalized Patients With 2019 Novel Coronavirus-Infected Pneumonia in Wuhan, China [published online ahead of print, 2020 Feb 7]. JAMA. 2020;323(11):1061–1069. doi:10.1001/jama.2020.1585.

[17] Huang C, Wang Y, Li X, Ren L, Zhao J, Hu Y, et al. Clinical features of patients infected with 2019 novel coronavirus in Wuhan, China [published correction appears in Lancet. 2020 Jan 30]. Lancet. 2020;395(10223):497–506. doi:10.1016/S0140-6736(20)30183-5.

[18] Richardson S, Hirsch JS, Narasimhan M, Crawford JM, McGinn T, Davidson KW, et al. Presenting Characteristics, Comorbidities, and Outcomes Among 5700 Patients Hospitalized With COVID-19 in the New York City Area [published online ahead of print, 2020 Apr 22]. JAMA. 2020; e206775. doi:10.1001/jama.2020.6775.

[19] Li Q, Guan X, Wu P, Wang X, Zhou L, Tong Y, et al. Early Transmission Dynamics in Wuhan, China, of Novel Coronavirus-Infected Pneumonia. N Engl J Med. 2020;382(13):1199–1207. doi:10.1056/NEJMoa2001316.

[20] Borges do Nascimento IJ, Cacic N, Abdulazeem HM, von Groote TC, Jayarajah U, Weerasekara I, et al. Novel Coronavirus Infection (COVID-19) in Humans: A Scoping Review and Meta-Analysis. J Clin Med. 2020;9(4): E941. Published 2020 Mar 30. doi:10.3390/jcm9040941.

[21] Guan WJ, Liang WH, Zhao Y, Liang HR, Chen ZS, Li YM, et al. Comorbidity and its impact on 1590 patients with Covid-19 in China: A Nationwide Analysis [published online ahead of print, 2020 Mar 26]. Eur Respir J. 2020;2000547. doi:10.1183/13993003.00547-2020.

[22] Li LQ, Huang T, Wang YQ, Wang ZP, Liang Y, Huang TB, et al. COVID-19 patients’ clinical characteristics, discharge rate, and fatality rate of meta-analysis [published online ahead of print, 2020 Mar 12]. J Med Virol. 2020;10.1002/jmv.25757. doi:10.1002/jmv.25757.

[23] Badawi A, Ryoo SG. Prevalence of comorbidities in the Middle East respiratory syndrome coronavirus (MERS-CoV): a systematic review and meta-analysis. Int J Infect Dis. 2016; 49:129–133. doi: 10.1016/j.ijid.2016.06.015.

[24] Gustafson PE. Gender differences in risk perception: theoretical and methodological perspectives. Risk Anal. 1998;18(6):805–811. doi:10.1023/b:rian.0000005926.03250.c0.

[25] Srivastav A, Santibanez TA, Lu PJ, Stringer MC, Dever JA, Bostwick M, et al. Preventive behaviors adults report using to avoid catching or spreading influenza, United States, 2015-16 influenza season. PLoS One. 2018;13(3):e0195085. Published 2018 Mar 30. doi: 10.1371/journal.pone.0195085.

[26] Fung IC, Cairncross S. How often do you wash your hands? A review of studies of hand-washing practices in the community during and after the SARS outbreak in 2003. Int J Environ Health Res. 2007;17(3):161–183. doi:10.1080/09603120701254276.

[27] Zhu Z, Cai T, Fan L, Lou K, Hua X, Huang Z, et al. Clinical value of immune-inflammatory parameters to assess the severity of coronavirus disease 2019 [published online ahead of print, 2020 Apr 22]. Int J Infect Dis. 2020; S1201-9712(20)30257-5. doi: 10.1016/j.ijid.2020.04.041.

